# Bilateral aldosterone production is most common in primary aldosteronism found by screening within primary care

**DOI:** 10.1101/2024.02.22.24303228

**Authors:** Nikita Makhnov, Jakob Skov, Fredrik Axling, Daniel Andernord, Mikael Bergenheim, Mauritz Waldén, Per Hellman

## Abstract

**OBJECTIVE:** Primary aldosteronism (PA) is the most common cause of secondary hypertension, and implies high risk of cardiovascular complications. Previously, our group had screened 1181 unselected Swedish primary care hypertensive patients, to find PA prevalence of 4,5%. The current study describes results of further diagnostics and specific treatment of these PA patients.

**DESIGN:** Prospective evaluation of lateralization and treatment of patients with PA.

**PATIENTS:** Fifty-three individuals with PA diagnosed by screening carried out in strict accordance with the current guidelines.

**MEASUREMENTS:** All patients accepting possible surgery underwent adrenal venous sampling, complemented, if unsuccessful, by adrenal cortex-specific positron emission tomography. Lateralizing cases were advised laparoscopic adrenalectomy. Bilateral ones were treated by mineralocorticoid receptor antagonists. Treatment results were assessed by at least 6 - 12 months of follow-up.

**RESULTS:** Localizing studies were accepted by 45 patients, 8 patients declined. Lateralized disease was found in 14 cases. Of 11 operated cases 3 had adenoma (prevalence 7-13%). Remaining operated cases comprised micro- and macronodular pathology. Thirty-one patients had bilateral PA. Screening and specific PA treatment led to better blood pressure and raising renin signifying risk amelioration. Both surgical and conservative treatment were tolerated well even when mainly conducted at regional healthcare level.

**CONCLUSIONS:** Bilateral production of aldosterone is common in a patient cohort derived from screening within primary care. Lateralization may be due to either adenoma, which is very rare, or due to nodular forms of PA, and both are successfully treated with adrenalectomy.

## Introduction

Primary aldosteronism (PA) may be defined as adrenal overproduction of aldosterone in excess of physiologic needs based on actual effective circulating volume, sodium and potassium status. It represents the most common cause of secondary hypertension ^1^ and is characterized by significantly worsened prognosis concerning cardiovascular and renal complications compared to primary hypertension. Due to frequent absence of specific clinical symptoms, the clue to diagnosis of PA lies in biochemical screening, most usually done by plasma aldosterone-renin ratio, ARR ^2^. Given the extent of arterial hypertension in general ^3^, and the fact that screening of hypertensive populations shows common occurrence of PA ^4^, it is of great importance to try to diagnose and adequately treat these patients. However, this strategy is counteracted by low awareness of the disease in the medical community ^5^, as well as by time consuming and expensive diagnostic and treatment algorithms ^2^. Research in this area is important as both more straightforward diagnostics and more efficient treatment strategies are needed.

Abnormal release of aldosterone from adrenal glands in PA may be uni- or bilateral. Historically, the unilateral cases were considered to be caused by an aldosterone-producing adenoma (APA), while bilateral cases were explained by bilateral hyperplasia. Progressive pathophysiologic research into the cellular nature of changes in the affected adrenal glands in bilateral and unilateral PA has recently resulted in the HISTALDO consensus on histological forms of the disease ^6^. The major part of the histopathologically studied adrenals has naturally come from patients operated for unilateral PA ^7 8^. Genuine, classical, hyperplasia according to HISTALDO criteria accounts probably for just a small fraction of unilateral non-APA forms of PA ^9 10^. Use of immunohistochemical analysis of the enzyme produced by *CYP11B2* (aldosterone synthase) have permitted to discern otherwise unseen overproducing micronodules, and even differentiate between nodules and adenomas seen by hematoxylin-eosin in relation to their aldosterone production. Thus, there is an emerging picture of many non-hyperplasia, in the classical meaning, and non-adenoma combinations of lesions which may cause unilateral PA. These lesions are comprised of macro- and/or micronodular areas, best revealed by CYP11B2 immunohistochemistry. Whether these are truly unilateral or asymmetrically bilaterally expressed in patients with lateralized PA is presently more or less unknown.

Our group has recently completed screening of 1181 hypertensive primary care patients according to the current guidelines, and diagnosed 53 of these individuals with PA - corresponding to a prevalence of 4,5%. The screening was done with ARR, followed by intravenous saline suppression test (SST) that, when necessary, was repeated after maximal clinically tolerated adjustment of medication potentially affecting the diagnostic results. The present study was aimed to describe localization studies and the following specific treatment of PA in this group. In addition, we noted the rates of adenoma and non-adenoma histopathological forms of disease in this cohort of hypertensive individuals.

## Materials and methods

### Study population

Fifty-three individuals recruited from the previous screening in a primary care setting all had confirmed PA. The study cohort is described in Tables 1 and 2, including medication before adjustments of treatment or lateralization procedures. The study was approved by the Swedish Ethical Review Authority, and registered on ClinicalTrials.gov (NCT03105531). Data protection laws were adhered to.

**Table 1:** Initial clinical features of the study group of 53 patients with primary aldosteronism (*median (interquartile range, IQR) or n (%))*

|  |  |
| --- | --- |
| Male | 27 (51 %) |
| Female | 26 (49 %) |
| Age (years): | 57(53-61) |
| Age when hypertension was discovered (years) | 44,5 (38-53) |
| Body mass index | 29,4 (25,9-33,1) |
| Systolic blood pressure | 145 (130 -153) |
| Diastolic blood pressure | 90 (85 – 95) |
| Serum creatinine (umol/l) | 73 (63 – 80) |
| eGFR (MDRD formula) ml/min <sup>a</sup> | 82 (75 – 90) |
| Serum sodium (mean (SD)) | 140,6 (1,83) |
| Serum potassium (mean (SD)) | 3,7 (0,32) |
| Former smoking | 21 (40 %) |
| Current smoking | 3 (6 %) |
| Former use of chewing tobacco | 12 (23 %) |
| Current use of chewing tobacco | 7 (13 %) |
| Hyperlipidemia <sup>b</sup> | 15 (28 %) |
| Diabetes mellitus or glucose intolerance | 8 (15,1 %) |
| Coronary artery disease <sup>c</sup> | 7 (13 %) |
| Sleep apnea syndrome | 7 (13 %) |
| History of arrhythmia <sup>d</sup> | 2 (4 %) |
| Known diagnosis of chronic renal failure | 1 (2 %) |
| History of stroke | 0 |
| Peripheral arterial disease | 0 |
| Known diagnosis of chronic heart failure | 0 |
| Chronic obstructive lung disease | 0 |
| History of pulmonary embolism | 0 |
| Osteoporosis | 0 |

<sup>a</sup> Estimated glomerular filtration rate, MDRD formula (Modification of Diet in Renal Disease Study Group)
<sup>b</sup> Hyperlipidemia was defined as chronic use of statins or other prescribed lipid-lowering medication
<sup>c</sup> Angina pectoris / myocardial infarction / percutaneous coronary intervention in the anamnesis
<sup>d</sup> Atrial fibrillation / flutter / other significant acquired chronic arrhythmia

**Table 2:** Initial number of groups of antihypertensive medicines in the study group of 53 patients with primary aldosteronism.

| number of groups: | n (%) | number of groups: mean (SD) | number of groups: median (IQR) |
| --- | --- | --- | --- |
| 0 | 4 (7,5 %) | 2,2 (1,28) | 2 (1-3) |
| 1 | 16 (30,2 %) |  |  |
| 2 | 9 (17 %) |  |  |
| 3 | 17 (32,1 %) |  |  |
| 4 | 5 (9,4 %) |  |  |
| 5 | 2 (3,8 %) |  |  |

### Diagnostic and treatment protocol

The flow chart for the study is represented in Figure 1. The group of patients with the diagnosis of PA (n = 53) were managed according to the Endocrine Society guidelines ^2^ and to the routine clinical practice in the region. It is worth mentioning that a considerable part of the studied PA cases had very mild clinical and biochemical signs of the disease.

**Figure 1.**
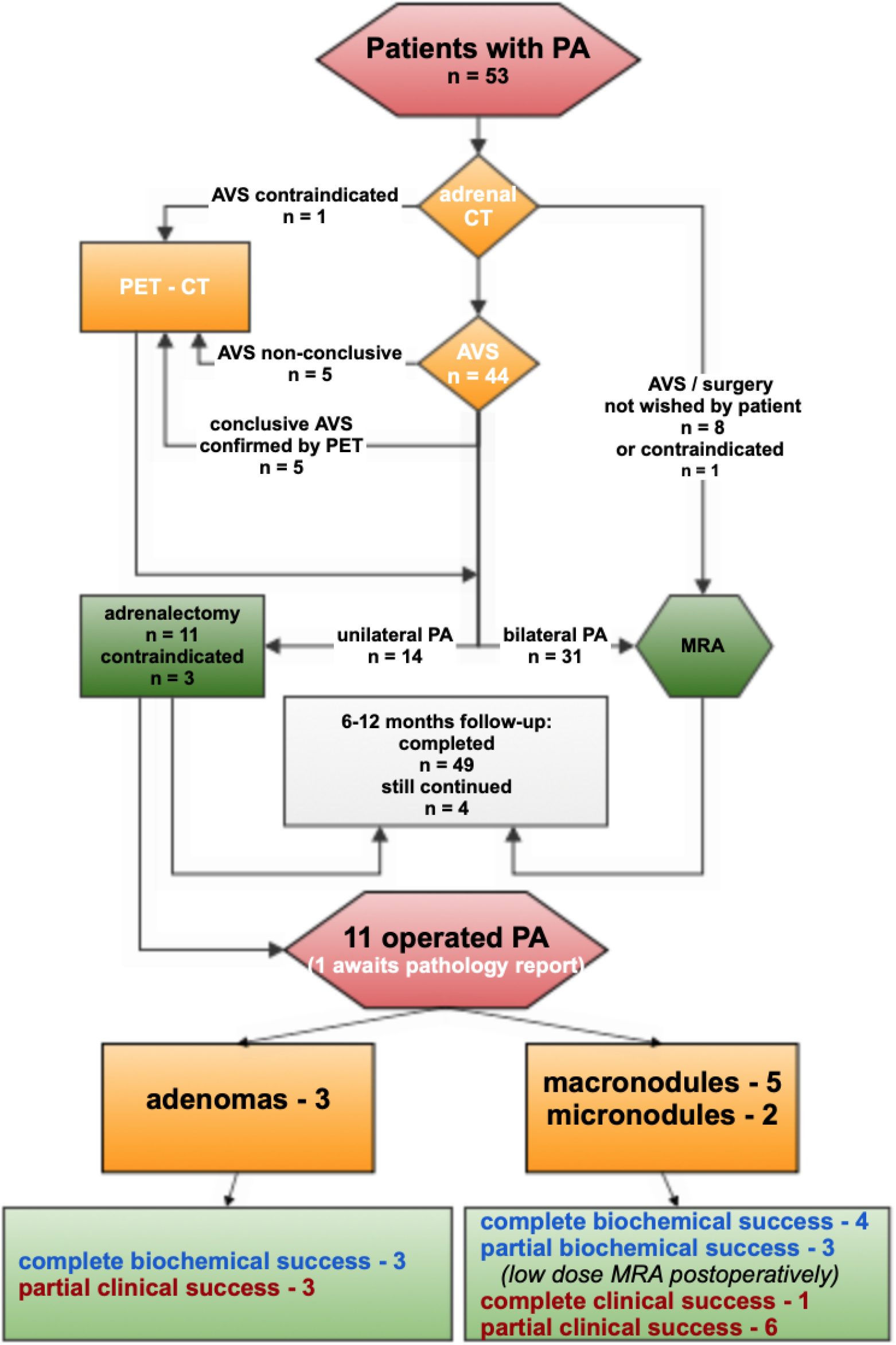
Work-up and treatment of 53 cases of primary aldosteronism; pathological, biochemical and clinical outcomes of operated cases. PA – primary aldosteronism; CT – computed tomography; AVS – adrenal venous sampling; PET-CT – positron emission tomography with computed tomography; MRA – mineralocorticoid receptor antagonists;

Computed tomography of adrenals without contrast was performed. Exclusion of cortisol- and catecholamine overproduction was assured with regular biochemical screening. Patients not objecting to the alternative of surgical treatment (n=44) were referred to Uppsala University Hospital for adrenal vein sampling (AVS) performed under current medication, but after pausing of mineralocorticoid receptor antagonists for at least six weeks.

AVS was performed without Synacthen stimulation, and was deemed successful if selectivity index (SI) on each side (cortisol in the respective adrenal vein divided by cortisol in the vena cava inferior) was ≥ 2,0. Aldosterone production was defined as dominant on one side if the lateralization index (LI, the greater ratio of aldosterone to cortisol in one adrenal vein divided by the smaller aldosterone to cortisol ratio in another adrenal vein) was ≥ 4,0. If LI was < 4 the PA was defined as bilateral. In one case, the inter-professional clinical decision was taken to recommend surgery to a patient with LI of 3,2 which was then effectuated.

In 11 patients adrenocortical positron emission tomography (PET/CT) was performed: in 5 where AVS was (sometimes repeatedly) not successful; in 5 – to confirm the result of successful AVS, and in 1 patient without prior AVS. Of these 11 PET/CT, 9 were done with 11C-metomidate (MTO) – that has been used as the adrenocortical PET tracer at Uppsala University Hospital for about 20 years ^11^, and the 2 most recent PET/CT were done with the newly adapted and somewhat more specific tracer para-chloro-2-[^18^F]fluoroethyletomidate (18F-CETO) ^12^.

The criterium for diagnosis of lateralized PA based on MTO- or CETO-PET/CT was the ratio of 1,25 between the (higher) SUV (standardized uptake value) in one adrenal divided by the (lower) SUV in the other adrenal. The ratio of 1,25 was chosen as it had been clinically used so by the Uppsala University Hospital’s PET-Center during a number of years preceding the current project – initially originating from the study of Burton et al. 2012 ^13^, that in retrospect even served as a basis for the later published MATCH-study ^14^. On clinical and pragmatic grounds concerning non-inferior sensitivity and specificity of CETO compared to MTO, the same ratio 1,25 was applied to cases subjected to CETO-PET/CT. However, the optimal ratio for 18F-CETO-PET is under ongoing investigation.

Patients with dominant aldosterone production on one side were advised laparoscopic unilateral adrenalectomy. Laparoscopic adrenalectomies were performed by the endocrine surgical team at the Surgical Department in Karlstad Central Hospital.

The patients who were not positive to surgical management (or those who were shown to have bilateral PA by AVS (LI<4 except in one case, see above) or by adrenocortical PET/CT) were started on slowly and progressively optimized doses of MRA (Eplerenone or Spironolactone) under repetitive control of blood pressure, plasma creatinine, sodium and potassium. The MRA doses were titrated with intention to have potassium within the higher normal range, and renin levels (at least 2 months after MRA start) no longer suppressed (at least above 8 mIU/L). Clinical follow-up after adrenalectomy or after MRA initiation continued within the study for at least 6 - 12 months.

In the present report, we also retrospectively noted the final results of the first confirming SST from the diagnostic phase, and related those to the lateralization data within the group.

## Statistical analysis

Variables were expressed as mean (standard deviation, SD) where parametric methods were employed, or as median (interquartile range, IQR) if nonparametric methods were used. Categorical variables were expressed as absolute numbers and percentages. Differences between groups with nonparametric testing were analyzed by Mann-Whitney U test or Wilcoxon signed-rank test. When appropriate, Students paired t-test was used. Categorical data were assessed by Fisher’s exact test. All the statistical tests used were two-sided. P ≤ 0,05 was considered significant. IBM SPSS Statistics version 26 program was used for statistical analysis.

## Results

### Lateralization of PA

Forty-four of 53 PA patients agreed to and underwent AVS (see Figure 1), permitting bilateral PA to be diagnosed in 28 cases and lateralized PA in 11. In 2 patients AVS was interpretable only when repeated the second time. In 5 cases the AVS was not interpretable due to technical difficulty of the catheterization and low SI (even in 2 of these 5 cases AVS was attempted twice); these 5 patients were subjected to adrenal specific PET/CT (showing 3 clearly bilateral and 2 lateralizing forms of PA). One patient underwent PET/CT without prior AVS, the uptake ratio signifying lateralized disease. Totally, 31% (14 out of 45) of all who underwent AVS or PET/CT demonstrated lateralization. There were no complications related to AVS or PET.

### Operated patients

Of the 14 lateralized cases, 11 have been operated with unilateral laparoscopic adrenalectomy (see Figure 1). Postoperative results were assessed using PASO- criteria ^15^. Postoperative histology showed 3 cases of adenoma and 7 cases corresponding to macro- or micronodules ^6^ (two of these 7 cases had “normal” postoperative hematoxylin-eosin histology, presented a complete biochemical success – and were thus presumed to correspond to the micronodular pathological form of PA). The 3 cases of adenoma all had complete biochemical success of surgery. The 7 cases of nodular pathology included 4 with complete and 3 with partial biochemical success results, and in the latter three patients low doses of MRA had to be initiated.

Complete clinical success (where the patient discontinued all antihypertensive medication postoperatively) was only seen in one case. This 56-year old female patient with a 25 year history of hypertension had micronodular lateralized PA.

Of all operated patients there was one case of postoperative pulmonary embolism in spite of pharmacological anti-thrombotic prophylaxis. Temporary cortisol substitution was needed in two cases - in spite of normal preoperative Dexametason suppressed cortisol (24 nmol/L respective 40 nmol/L). There were otherwise no postoperative complications.

### Low proportion of aldosterone producing adenomas (APA)

Only 3 of the operated patients had an adrenocortical adenoma in the specimen, and 3 of the unilateral PA-patients have not been operated. Thus, 3-6 out of 45 individuals who underwent localizing procedures had adenomas (6,7-13,3%), and among those who lateralized - 3-6 out of 14 had adenomas (21,4-42,8%). Consequently, 8-11 out of 14 who lateralized has some form of true hyperplasia or nodular disease (57,1-78,6%). Assuming that the 31 who did not lateralize also had some form of non-adenoma disease, the total number of patients with non-adenoma disease finally reaches 39-42 out of 45 (86,7-93,3%).

### Conservatively treated PA

Forty-two of the PA patients: all of the 31 patients with the bilateral forms (AVS 28 + MTO 2 + CETO 1), 8 who refrained from AVS, and 3 unilateral (2 AVS + 1 PET) (who finally had to be treated conservatively) were started on MRA with the intention to follow treatment until the MRA dose was considered to provide adequate elevation of plasma renin while keeping serum potassium within higher normal reference range. Thus, this subcohort consisted of patients with both lateralized and non-lateralized disease. Within a median of 13 (IQR 12-17) months, 40 of these patients (40 out of 42; 95,2%) were considered to have reached the optimal possible MRA therapy. Conservative treatment was chosen for the three lateralized PA-patients as: one of them had a gigantic adrenal cyst with pressure-symptoms demanding adrenalectomy, which turned out to be on the opposite side to the aldosterone overproducing adrenal; the other patient was lateralized by PET/CT but deemed to not qualify for surgery due to morbid obesity; the third patient decided already after AVS to decline surgery in view of then already well functioning MRA treatment.

The three cases with partial biochemical success postoperatively (all with non-adenoma forms of aldosterone overproduction) were initiated on low doses of MRA and responded with adequate rise in renin.

**Protective raised level of renin at the completion of the study participation** was achieved by the absolute majority of the patients with PA. The direct renin concentration (DRC) value of 8 mIU/L corresponds to minimal protective plasma renin activity (PRA) value of 1 ug/L per h ^16^ – even if the cited study strictly concerned conservatively treated PA patients.

Final DRC was available in 47 of 49 patients who completed the follow-up, and was > 8,0 mIU/L in 42 of these 47 patients (89,4 %). In 2 patients with complete biochemical success after adrenalectomy, demonstrated as postoperatively low ARR and PAC at a lower normal level, DRC below 8 mIU/L was still present. In addition, in 3 of the conservatively treated patients final DRC was below 8 mIU/L – but in 2 of them this was deemed due to concurrent administration of beta blockers and nonsteroidal anti-inflammatory drugs (NSAID), and one patient with totally normalized blood pressure on a given dose of MRA wished not to raise the MRA dose further– in spite of somewhat low final renin. Just one patient (with ongoing NSAID treatment) had final DRC slightly below its formal lower reference range, the rest of the above cases had DRC above the formal lower reference level of 4,4 mIU/L for the upright position.

As retrospectively noted in these 47 patients, the last DRC before the initial saline suppression test was median 4,1 (IQR 1,8 – 5,7) mIU/L. Final study DRC for both conservatively treated and operated patients was 20 (9,8 – 44) mIU/l, thus being significantly higher (Wilcoxon signed-rank test, p < 0,001).

### Treatment-related effect on serum potassium

Of 53 PA cases, 23 had to have potassium supplementation before screening with ARR. None of the patients with finally established treatment of PA needed potassium preparations any longer (Fishers exact test p < 0,0001). As expected, potassium levels within the group of PA (data is available on n=46) were significantly higher after adequate treatment (mean 4,26 (SD 0,28) mmol/L) than initially before screening (3,73(0,32)); p < 0,001.

### Study-related effect on use of blood pressure medication

The number of groups of antihypertensive medications initially at inclusion and after completed PA treatment was available for 49 patients. The ambition during the final adjustment of medication was to achieve as normal blood pressure as possible. That may well lay behind the result that the number of blood pressure medications rose slightly for the study patients (the mean rose from 2,16 before the study to 2,57 at its conclusion). Before the study, the number was median 2 (IQR 1-3), and at the completion of the study 2 (2-4). Wilcoxon signed-rank test gave p = 0,034.

**Positive effect of study-related treatment on blood pressure** was noted, see Table 3. The distribution of the differences between systolic / diastolic pressure before and after application of specific PA treatment (data is available for 47 patients) included 1 outlier. After its exclusion, a paired t-test was possible – but paired t-test including the outlier produced basically the same result (and the same high degree of significance of differences), why the results are shown for all of the followed-up patients. Substantial improvement of blood pressure was achieved by treatment. Just one patient had final systolic pressure slightly above 140, and all patients had diastolic pressure below 90 by the end of the follow-up period of median 12 (IQR 12-17, range 6-33) months.

**Table 3:**
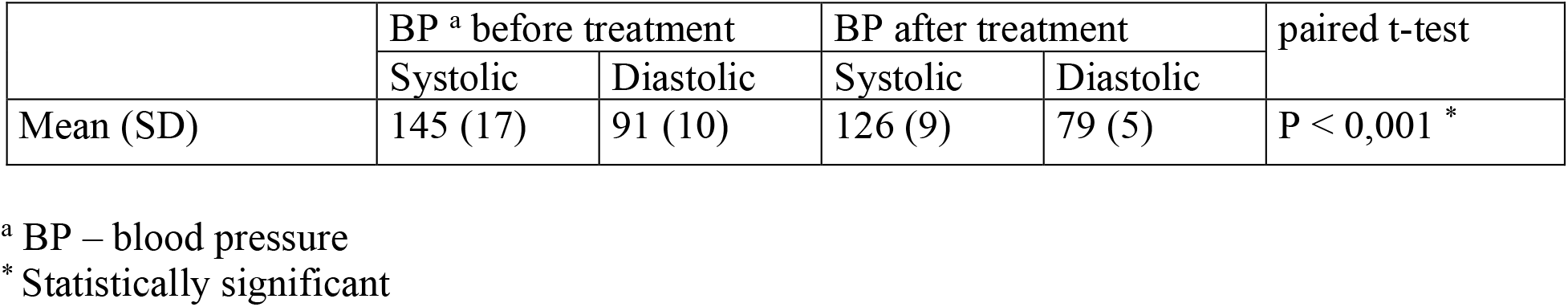
Specific treatment of primary aldosteronism: effect on blood pressure (n = 47)

### Higher aldosterone production in unilateral cases

Final aldosterone level after initial diagnostic SST in the group with localized PA was 320 (270-464) pmol/L, significantly higher than in the group with bilateral PA (246 (203-300) pmol/L). Mann-Whitney U test gave p=0,009.

## Discussion

The majority of published studies, guidelines and educational sources to date adhere to the longstanding tradition of dividing the anatomical conditions behind PA in “unilateral aldosterone producing adenoma (APA), representing 30-40 % of cases”, “bilateral hyperplasia, representing 60-70 % of cases” and other seldom occurring forms of PA (sometimes mentioning “unilateral hyperplasia”).

Lately, parallel to advances in screening for PA leading to its milder forms being recognized, there appeared a number of reports of “unilateral hyperplasia” as an operable cause of the disease. One of the first publications appeared 2002 ^17^, an early review came 2007 ^18^, and a comprehensive study about the role of non-adenoma cause for unilateral PA was published 2012 ^19^ – where Iacobone et al. showed that a considerable part of all cases of lateralized PA were represented by non-adenoma pathology.

There is a need for a linguistic commentary here due to the mentioned HISTALDO consensus and its recommended nomenclature ^6^. The basis for this classification is derived from the immunohistochemical (IHC) localization of expression of aldosterone synthase (CYP11B2) in the adrenal cortex. Adenoma (by definition at least 10 mm i maximal diameter), visible in hematoxylin-eosin (HE) staining, may thus not show CYP11B2 and then be defined as non-functioning. Single or multiple macronodules (less then 10 mm, but seen both by IHC and HE) and micronodules (by definition seen only on IHC but not in HT staining) comprise a big part of found lesions. Unilateral diffuse hyperplasia positive for CYP11B2 is one of the more rare findings ^10^ - and thus a traditional inclusive term of “unilateral hyperplasia” is proposed to lose its wider sense. Iacobone et al. have seen that up to 74% of AVS defined unilateral PA cases were represented by non-adenoma lesions (even if IHC was not performed), and their study results for unilateral adrenalectomy in these cases were as highly effective as in those due to APA (based on a median of 3 years of follow-up). Of totally 26 non-adenoma lesions, 23 included macronodules, and 3 were described as hyperplasia seen in HE-staining.

Most likely can these “unilateral” non-adenomatous cases be retrospectively, in HISTALDO terms, seen as expressing macro- and micronodules. For obvious reasons, it is unknown whether the contralateral adrenal glands also expressed the same structures but in low quantities. The study’s relatively short follow-up does not permit further speculation.

Unilateral non-adenoma disease in that study reached at 6 months follow-up as good cure after adrenalectomy as did the cases caused by adenoma. Whether even the contralateral gland would possibly expresses nodular pathology can only be seen if an eventual recurrence of aldosterone excess is revealed by long-term follow-up, and even then would it be unlikely to get a histologic description of the remaining adrenal gland.

Diagnosis and treatment of PA has traditionally and unavoidably been associated with a set of issues that still make it difficult on any national level to cite better results than a few percent of cases discovered and appropriately managed ^20 21 5^. The reason lies undoubtedly partly in a level of intensity of procedures in each given case to come to a definitive diagnosis, partly in an invasive manner of the work-up necessary to be able to offer surgery to unilateral cases.

Besides, therapy with MRA is also in the long run a cumbersome process necessitating regular controls of potassium, creatinine, and consideration of interactions with medications that can lead to lowering glomerular filtration rate or raising potassium levels. Traditional excessive caution of the primary care practitioners and even internal medicine specialists has in fact been preventing patients from reaching sufficient doses of MRA that would effectively protect from consequences of PA. Surgical treatment (of unilateral forms of PA) is nowadays connected with the best long term risk reduction for these patients ^22^, at the same time as even specific and continued pharmacological treatment (especially in those cases when it is not sufficient enough to counteract renin suppression) is associated with significantly greater secondary organ damage and increased mortality ^16 23 24^.

Hundemer et al. ^16^ found in a retrospective cohort study (comparing 602 MRA-treated PA- patients with matched HT-cases) hazard ratios for PA-cases of 2,8 for cardiovascular complications, and of 1,8 for death - if their plasma renin activity (PRA) was under 1 μg/L per h. PA-patients on MRA with PRA ≥ 1 μg/L per h had a risk profile similar to those with primary hypertension.

The role of screening for PA in a hypertensive population is thus not just to help discover the condition and improve control of blood pressure: it is as important to adequately reduce the effects of non-physiologic aldosterone levels.

Lateralized PA is generally characterized by more pronounced aldosterone overproduction than non-lateralized forms – which may permit limiting the number of patients for localizing studies as those are essentially needed just for the potentially operable cases. ^25 26^ This paradigm is even illustrated in our material where final aldosterone level after SST in the diagnostic phase was significantly higher among the unilateral cases compared to the bilateral cases.

Naturally, there are some patient groups (especially those with mild forms of PA) where localizing studies would not be clinically warranted ^27^. Conservative treatment in such individuals, together with further development of non-invasive localizing methods such as adrenal cortex-specific PET/CT, can hopefully raise possibility to give adequate specific treatment to a bigger part of patients with PA and improve accessibility for localizing diagnostics to those with probable lateralizing disease. Even in our present material, a majority of patients had clinically and biochemically mild PA (probably as the study group came from screening of an unselected population of primary care hypertensive patients), and a majority of patients demonstrated bilateral disease. Obviously though, for the time being, there are no established non-visualizing methods to effectively define the group with high prevalence of a localized form of PA.

High prevalence of nodal forms and relatively low prevalence of APA may be a feature of the lateralizing PA cases found in a setting of liberal screening within primary care hypertensive patients.

## Limitations

An obvious limitation of the study is a modest number of patients with PA. The fact that they coherently originate from an unselected primary care hypertensive population may on the other hand be seen as a strong advantage - thus detailing the profile of PA that may be the one more typical for the general majority of the cases.

## Conclusion

The report demonstrates that liberal application of current guidelines in diagnostics of hypertensive primary care patients results in a cohort of patients with PA expressing fewer adenomas (APAs) than regularly noted in the literature. This signifies a need for change in the histopathological vision of the disease where the traditional classification of cases in “unilateral with APA and bilateral with hyperplasia” may not mirror the reality among relatively mild forms of the condition. We also describe the use of adrenocortical PET/CT among the localizing studies, and foresee an increased role for this non-invasive method in the future.

Clinical work within the study (including all the aspects of treatment) was carried out in a regional hospital (with AVS and PET done at the university hospital). It is not improbable that the scope of PA diagnosis in adult population one day reaches quite considerable volumes. In that case, the study may serve as an example of feasibility of decentralization of practical treatment for PA.

## Data Availability

Research data are not shared due to privacy or ethical restrictions.

## Acknowledgements

The skillful and dedicated assistance by the project nurse Maria Nordström is greatly appreciated. The authors also express their gratitude to all the clinical and administrative staff who helped and supported the project.

